# Vaccine Approach for Human Monkeypox over the Years and Current Recommendations to Prevent the Outbreak: A Rapid Review

**DOI:** 10.1101/2022.09.29.22280481

**Authors:** Rifat Ara, Tajrin Rahman, Rima Nath, A.M. Khairul Islam, Miah MD Akiful Haque, Md. Ferdous Rahman, Mohammad Hayatun Nabi, Mohammad Delwer Hossain Hawlader

## Abstract

World Health Organization has declared human monkeypox as a global health emergency on 23 July 2022. This indicates that the outbreak poses a serious risk to global health and requires a united worldwide response to stop the virus from spreading and possibly turning into a pandemic. Vaccines can play a vital role in this context, contributing to pre- and post-exposure prophylaxis. The aim of our rapid review was to go through the background of the vaccine approach for human monkeypox over the years and to find out what current guidelines are highlighting relating to it. 22 relevant published articles from MEDLINE bibliographic database and 8 vaccine recommendations from manual searching have been deliberated here. The significant synopsis of this review is that the smallpox vaccine is the only immunization option for monkeypox so far, and it is up to 85% effective to prevent the infection. Third-generation smallpox vaccines are advised over first and second generations due to their minimal side effects. Healthcare providers and lab professionals at risk are on the priority list to get vaccinated, as well as pregnant women or lactating mothers, immunocompromised or chronic ill patients, can get vaccinated if they are surely exposed to the monkeypox infection. Lastly, JYNNEOS/IMVAMUNE is the current most preferable smallpox vaccine that is highly advised for the latest outbreak of human monkeypox but more clinical trial on humans should be conducted to evaluate its safety, efficacy, and adverse events.

## Introduction

Orthopoxviruses belong to the Poxviridae, the family of enclosed viruses with large linear, twofold DNA genomes (0.130 kb genome) [1]. Multiple species of this group can cause serious human illness. Variola virus is the major human pathogen in the family Orthopoxvirus and the causal agent of smallpox. Concerns have been expressed over the use of the variola virus or a similar genetic pathogenic orthopoxvirus as a bioweapon [2,3] despite the elimination of smallpox. In addition, there is fear that the variola virus could be discharged inadvertently, such as from unused viral stocks. Variola virus was discovered cold-stored in a United States research laboratory, lending validity to the second possibility [4]. Orthopoxviruses, such as monkeypox virus, cowpox virus, and strains of vaccinia virus, are emerging zoonoses in many parts of the world [5–7]. These viruses cause significant diseases in both people and livestock. In reality, both the frequency and severity of human monkeypox virus epidemics have increased [6,8,9]. Monkeypox (MPX) is an infectious zoonotic disease largely concealed throughout the smallpox era. It was not recognized as a human disease until the late stages of smallpox eradication campaigns. MPX’s concealment might be linked to the comparable clinical manifestations of monkeypox and conventional smallpox, making it difficult to distinguish between the two. How the natural monkeypox virus is sustained in the wild is a significant outstanding question. Currently, natural monkeypox is restricted to humid forest regions in West and Central Africa [10]. Nevertheless, the disease incidence in the Democratic Republic of Congo has increased since 2001 [6,11]. The 2017–18 outbreak of monkeypox in Nigeria spurred the launch of a comprehensive surveillance strategy. Therefore, it is necessary to continue developing and refining orthopoxvirus countermeasures [12].

Early vaccinations used to eliminate smallpox used live, unattenuated vaccinia virus strains derived from calf lymph, like Dryvax [13]. These vaccinations are no longer manufactured and were replaced by ACAM2000 [14], a cultured cell, live vaccinia virus vaccine, and MVA (Modified Vaccinia Ankara), a replication-deficient (in human tissues) vaccine. Both calf lymph and cell culture vaccinations can cause severe adverse reactions in humans, involving autoinoculation of an eye, widespread vaccinia, eczema vaccinatum, recurrent vaccinia, myocarditis, as well as death [13,15,16]. In the absence of a more significant global threat posed by orthopoxvirus illness, these safety issues make the broad usage of this vaccine unethical. A substantial number of individuals are inappropriate for ACAM2000 due to security concerns, particularly those with immunological weaknesses and common skin disorders such as dermatitis [14]. ACAM2000’s safety issues encouraged the development of more extensively attenuated third-generation vaccines, such as MVA and Lc16m8 [17]. The U.S. Food and Drug Administration has recently approved JYNNEOS, the Barvarian Nordic variant of the MVA vaccine, to prevent smallpox. JYNNEOS protects animals from lethal orthopoxvirus infection, including monkeypox virus disease of nonhuman primates, rabbitpox virus infection of rabbits, as well as vaccinia virus infection of mice [18,19]. According to statistics from Africa, the smallpox vaccine is at least 85% effective at preventing monkeypox [20]. Similar to ACAM2000, MVA lacks known protective targets, and both infections express thousands of genetic variants unlikely to assist in protection [21].

### Current outbreak of monkeypox

Few occurrences of monkeypox outbreaks in humans have been reported outside of the African continent since the 2003 outbreak in 11 states of US [22]. After that a minor uptick has been observed in 2017 but the outbreak of 2022 illustrates a distinct situation that may have been predicted because there were early indicators [23]. 3413 cases with laboratory confirmation and one death have been reported to WHO from 50 countries in five WHO Regions since January 1 as of June 22, 2022. 86% of cases with test confirmation were reported from the WHO European Region [24]. As of 23^rd^ July 2022, WHO has issued the strongest call to action, it is able to in relation to the global monkeypox outbreak by declaring it as a public health emergency of international significance [25].

### Objective of this review

Several vaccine approaches have been applied over the years to prevent the spread of monkeypox infections. A vaccine has recently been approved for monkeypox; however, it is not yet widely available [26]. Some countries may have smallpox vaccination products on hand that might be used if national guidelines are followed. Depending on the country, any request for vaccination items may be offered in limited numbers through federal authorities. Governments may want to explore immunizing close contacts as a post-exposure prophylactic measure or immunizing select groups of healthcare personnel as a preventative measure. Considering the recent clustered outbreak of monkeypox in several countries and declaring it as a global public health emergency, this rapid review aims to explore different vaccine approaches and plan over the years, challenges and current recommendations to prevent the outbreak.

## Methodology

To conduct we this rapid review, searched MEDLINE bibliographic database and relevant websites to identify potential articles describing several vaccine strategies, vaccine efficacy, and vaccine challenges to prevent the monkeypox outbreaks. A simple search term “Monkeypox” AND “Vaccine” was used to find out all the pertinent write-ups, and all published documents up to the date of 30 June 2022 were included. Rayyan QCRI tool has been used to screen the articles. Primarily we retrieved a total of 291 articles, and duplicate articles (n= 4) were excluded. Two reviewers scrutinized 115 out of 287 articles through independent dual screening of titles and abstracts. Finally, 22 articles were selected after full-text evaluation, and data extraction was considered. PRISMA flow diagram describes the detailed procedure of article selection **(Figure 1**). In the case of selecting articles, the availability of the information regarding different vaccine approaches for monkeypox, and their outcome were under primary consideration. Besides, vaccine complications and challenges were our secondary objective to include in our review. This review included all relevant published documents, regardless of study type or geographical distribution, such as original articles, perspective papers, review papers, workshop reports, editorial letters, or comments. Because a translator was not available in the team, articles written in languages other than English were excluded.

**Figure 1:**
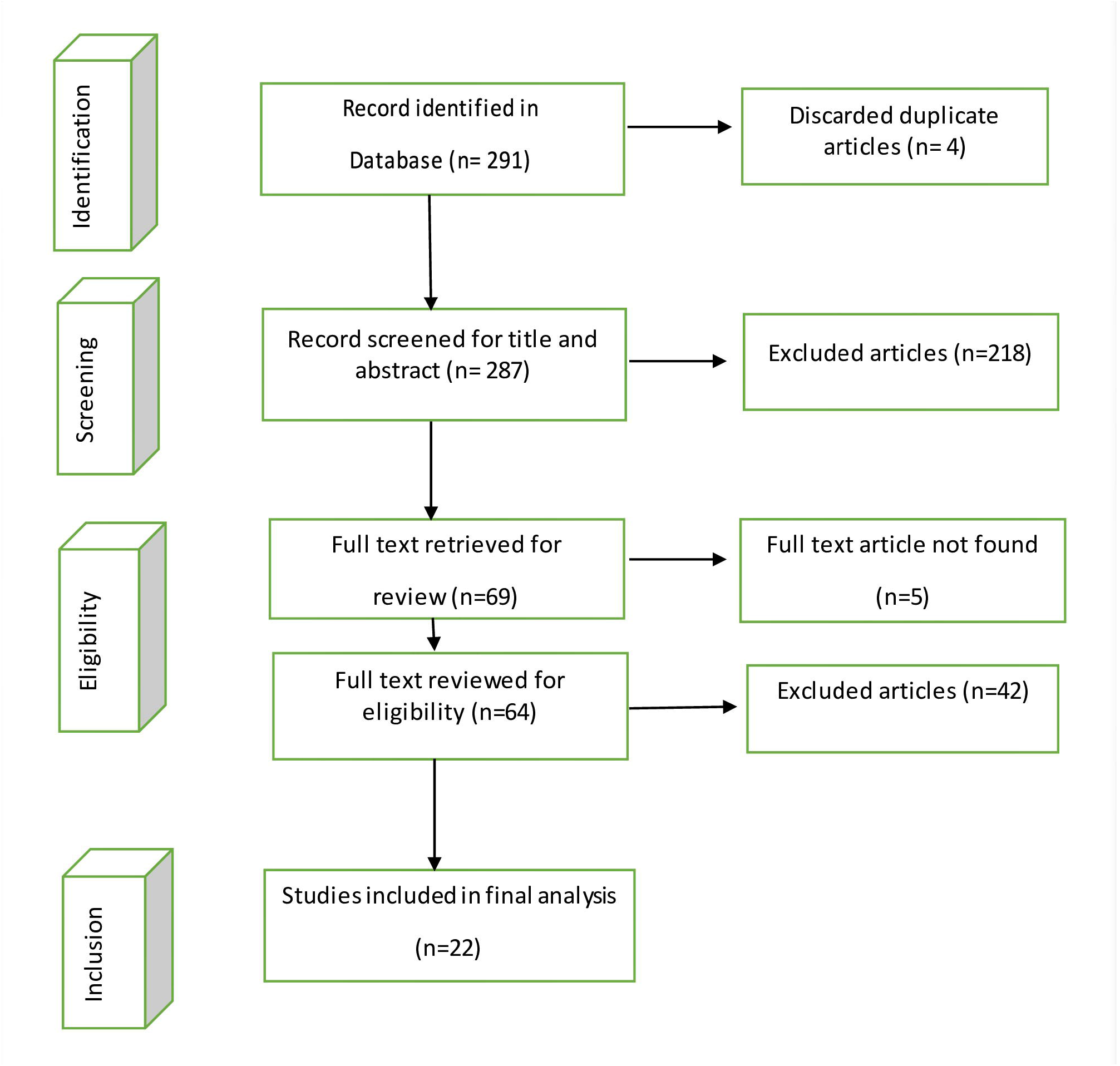
PRISMA flow diagram for article screening and selection.

For data extraction, key points; such as article types, publication year, study design, countries where vaccines have been experimented with, targeted population, name and types of vaccines, their clinical outcome (e.g., efficacy, effectiveness), challenges for vaccine actualization were assembled and recorded in a structured excel format. Moreover, we also followed the recommendations mentioned in the selected articles. We also did manual searching along with the literature review to find out current recommendations by different international health organizations regarding the Monkeypox vaccine guidelines. We mainly focused on the advice from the World Health Organization (WHO), Centers for Disease Control and Prevention (CDC), United Nations, UK Health Agency Security, Germany’s Standing Committee on Vaccination (STIKO), Gavi- the vaccine alliance, and Canada’s National Advisory Committee on Immunization (NACI). Recent vaccine recommendations, guidelines, and other information were gathered to map out the updated status of Monkeypox vaccines.

## Discussion

### Vaccine approach for human monkeypox over the years

A total of 22 articles have been found that analyzed and described the monkeypox preventive approaches through vaccination (Table 1). Over the years, researchers have explored active and passive surveillance, cohorts, public health investigations, and epidemiological data on monkeypox outbreaks and smallpox vaccination to see the efficacy and effectiveness of different generation vaccines against monkeypox infection. As a result, articles published from 1985 to date were included in our review. Our review perceived that all the articles talked about routine smallpox vaccines of different generations, and no specific vaccine has been manufactured for the monkeypox virus until now.

There are five articles published in 1980s that mainly investigated individuals of the African region who were routinely vaccinated against smallpox during their childhood and later exposed to the monkeypox virus. Among these five articles, Arita I. et al. assessed the special surveillance of tropical rain forests of West and Central Africa between 1970 to 1981 and 1982 to 1983, where a 13% case-fatality rate was achieved, and all of them happened to unvaccinated youngsters. Moreover, the secondary attack rate in unvaccinated contacts was around 15% [27]. The other four articles analyzed data on the monkeypox outbreak in Zaire of the Democratic Republic of Congo (DRC) over five years (1980-1984). They concluded that the standard routine smallpox vaccine was 85% effective in preventing monkeypox [28], and the secondary attack rates were 0.110 in unvaccinated contacts living in the same households [29,30].

After the 1980s outbreak of monkeypox in African territories, a long break took place studying preventive measures for monkeypox until the outbreak emerged in the USA in 2003. The first indication of community-acquired monkeypox in the United States was reported by the Centers for Disease Control and Prevention (CDC) at the beginning of June 2003, and CDC advised the vaccinia smallpox vaccine as it was established to be 85% effective against monkeypox [31]. Seven articles have discussed the effectiveness of past smallpox vaccination among US citizens to prevent monkeypox, where almost all the studies found positive outcomes of vaccination except one. A retrospective analysis of clinical reports and active and passive surveillance of suspected monkeypox cases was done in 2005, where bivariate and multivariate analysis found no association of disease severity or hospitalization with previous smallpox vaccination [32]. Vaccinated subjects showed good protection and less pronounced clinical sign symptoms against the onset of Monkeypox-induced disease in the rest of the studies [33-35]. Another significant public health investigation was done in 2005, where 94% of the monkeypox-exposed healthcare workers tested positive for anti-orthopoxvirus IgG antibodies as they had a previous history of smallpox vaccination [35]. Jamieson D.J. et al. and Cono J. et al. are the two articles that put forward the opinion regarding smallpox vaccination during pregnancy [31,36]. According to them, the smallpox vaccine is considered category C (the human fetal risk of the drug as unknown due to no human studies or positive results in animal studies) approved by the US Food and Drug Administration (FDA), and minor risk of fetal from vaccinia smallpox immunization during pregnancy exists that may result in premature birth, fetal and neonatal death. Nevertheless, given the potentially fatal risk of monkeypox infection, exposed women were urged to get the smallpox vaccine regardless of whether they were pregnant.

Another significant finding of our review was that despite having multiple animal trials of the vaccine directly against monkeypox, there is no sufficient evidence of clinical trials on humans yet. In recent times, a phase I/II randomized, double-blind, comparative clinical trial of LC16m8 (an attenuated cell culture–adapted Lister vaccinia smallpox vaccine) has been conducted on humans to compare the safety and immunogenicity of LC16m8 with the Dryvax vaccine [37]. This trial showed that LC16m8 is a feasible next-generation vaccination alternative to first-generation smallpox vaccines to prevent human monkeypox, at least in high-risk groups. Although, its clinical efficacy against human monkeypox has not yet been determined. A clinical trial of a new generation IMVAMUNE® smallpox vaccine is ongoing to evaluate its safety and efficacy in preventing monkeypox among HCWs in the Democratic Republic of the Congo [38].

Right now, the Strategic National Stockpile (SNS) has three smallpox vaccines, among which ACAM2000® and JYNNEOSTM (also known as IMVAMUNE or IMVANEX) are the only two licensed smallpox vaccines in the United States [39]. It has been found that the only smallpox vaccination effective in people of DRC was live vaccine inoculation (Dryvax and ACAM2000) [40]. Monkeypox risk was 5.21 times lower in vaccinated individuals compared to unvaccinated individuals, showing that more than 80% protection was maintained for over 30 years [40]. A prospective cohort study was performed in 2019 to follow up the health workers of DRC who have received two doses of the third-generation smallpox vaccine (IMVAMUNE) due to reporting so many adverse events of first- and second-generation smallpox vaccine [41]. Third-generation smallpox vaccine was better than the first- and second-generation. Though a clinical trial is ongoing to evaluate the safety and efficacy of the IMVAMUNE vaccine against human monkeypox among healthcare workers, an online based cross-sectional study was conducted among Indonesian health workers to evaluate the acceptance and willingness to pay (WTP) for the vaccine, where 96% of the participants expressed the acceptance of free vaccination [42]. A case study in Singapore revealed that 64% of close contacts recovered rapidly from sign symptoms of monkeypox due to accepting the ACAM2000 vaccine (Sanofi Pasteur Biologics Co) immediately after contact tracing [43].

The rate is inferior if we look into the recent smallpox vaccination status. In order to prevent smallpox from reemerging, WHO kept a stockpile of 200 million doses in 1980. However, when smallpox did not resurface in the late 1980s, 99% of the stockpile was destroyed [44]. By 2019, the United States had received 269 million doses of ACAM2000 and 28 million doses of MVA [45,46], but by the time the 2022 monkeypox outbreak began, only 100 million doses of ACAM2000 and 65,000 doses of MVA remained in the stockpile [47]. One of our included articles showed that only 10.1% of Nigeria’s population had received the smallpox vaccine as of 2016, and the serologic immunity level was 25.7% among those who had received the vaccination compared to 2.6% in the general population [48]. However, worldwide vaccination data is not known yet. Finally, there is one article that criticized the cost-benefit of mass vaccination. Bankuru S.V. et al. describe a compartmental epidemiological model and game theory approach that evaluated vaccination decision-making of a community [49]. The model quantifies the smallpox vaccine’s costs and advantages. This study determined that the ideal vaccination rate is approximately 0.04, meaning people should get vaccinated once every 25 years. Additionally, they discovered that monkeypox is preventable and can be eliminated through vaccination in a semi-endemic equilibrium (an infection that spreads only part of the year in a specific area). However, vaccination alone cannot wholly eradicate monkeypox in an equilibrium where it is entirely endemic [49].

### What is the latest vaccine recommendation for human monkeypox?

After manual searching for vaccine recommendations, several guidelines were put in place suggested by different international health organizations (Table 2). According to WHO, mass vaccination for monkeypox is not required so far. They also advise administering a suitable second or third-generation smallpox vaccine to contacts as post-exposure prophylaxis (ideally within four days of initial exposure) and healthcare workers at risk as pre-exposure prophylaxis [50]. CDC recommends JYNNEOS™ for certain laboratory workers and clinic teams who are susceptible to virus exposure. JYNNEOS™ is usually in two doses, given four weeks apart. People who have received other types of smallpox vaccine in the past might be considered for one dose only. Booster doses are recommended every 2 or 10 years if a person remains at continued risk for exposure to orthopoxviruses. JYNNEOS™ is recommended for individuals exposed to the monkeypox virus regardless of concurrent illnesses, pregnancy, breastfeeding, or poor immune system [51]. On the other hand, CDC also advised ACAM2000 immunization for military personnel and lab workers only, and they are not suggested during immunocompromised conditions as ACAM2000 has the potential for more side effects and adverse events than JYNNEOS [52,53]. The Vaccine Alliance-GAVI has recommended the vaccinia smallpox vaccine over the first generation and suggested increasing the availability worldwide due to the monkeypox outbreak [54]. Other national health organizations, such as the National Advisory Committee on Immunization (NACI)-Canada, the U.S. Department of Health and Human Services (HHS), and Germany’s Standing Committee on Vaccination (STIKO), have also suggested the latest smallpox vaccine (JYNNEOS/IMVAMUNE/IMVANEX) to fight against the recent outbreak of human monkeypox [55-57].

## Conclusion

As the rapidly spreading monkeypox outbreak in 2022 represents a global health emergency, the WHO labeled it as a “public health emergency of international concern (PHEIC)” and asked for an international response to collaborate on sharing vaccines and treatments [58]. So, governments worldwide should come forward immediately to impose preventive measures, including screening, isolation, and vaccine prophylaxis where necessary.

The limitation of our review was that we only searched the MEDLINE database due to the target of rapid review within a short period of time. A systematic review can also be performed in this regard, especially on vaccine clinical trials. Despite having some deficiencies, this rapid review on vaccine approach and recommendations concluded with several significant points, including-

- The smallpox vaccine has been the only immunization option for human monkeypox till now. There is no specific vaccine manufactured only for monkeypox yet.
- The smallpox vaccine is up to 85% effective in preventing monkeypox infection and it can provide protection for a very long period.
- Three categories of smallpox vaccine are there. The first-generation vaccine is not available in the market anymore. The third-generation vaccine is recommended over the first and second generation due to fewer side effects and adverse events.
- Smallpox vaccine is recommended for all monkeypox virus exposed individuals regardless of pregnancy, chronic illnesses, or poor immunological conditions.
- JYNNEOS/IMVAMUNE is the latest third-generation smallpox vaccine that almost all international health organizations have mostly recommended. A human clinical trial of this vaccine is currently ongoing, information which would be very much helpful to evaluate the safety and efficacy of the vaccine. More trial studies should be conducted in the future to find out the accuracy.
- The vaccine is highly recommended as pre-exposure prophylaxis to all healthcare workers at risk and contact as post-exposure prophylaxis, but mass vaccination is not required until now.
- Additionally, human monkeypox is preventable and can be eradicated through vaccination alone in a semi-endemic equilibrium but not in a fully endemic equilibrium.

## Ethical Considerations

The approval for this rapid review has been taken from Ethics Review Committee of North South University, Bangladesh. This study will not directly involve any human participant; therefore, consent will not be required.

## Data Availability

All data produced in the present work are contained in the manuscript.

## Acknowledgement

None

## References

1. Moss, B. Smallpox Vaccines: Targets of Protective Immunity. Immunol. Rev. 2011, 239, 8–26, doi:10.1111/J.1600-065X.2010.00975.X.

2. Smith, G.L.; McFadden, G. Smallpox: Anything to Declare? Nat. Rev. Immunol. 2002, 2, 521–527, doi:10.1038/nri845.

3. Noyce, R.S.; Lederman, S.; Evans, D.H. Construction of an Infectious Horsepox Virus Vaccine from Chemically Synthesized DNA Fragments. PLoS One 2018, 13, e0188453, doi:10.1371/JOURNAL.PONE.0188453.

4. Reardon, S. Forgotten NIH Smallpox Virus Languishes on Death Row. Nature 2014, 514, 544, doi:10.1038/514544A.

5. Essbauer, S.; Pfeffer, M.; Meyer, H. Zoonotic Poxviruses. Vet. Microbiol. 2010, 140, 229–236, doi:10.1016/j.vetmic.2009.08.026.

6. Rimoin, A.W.; Mulembakani, P.M.; Johnston, S.C.; Lloyd Smith, J.O.; Kisalu, N.K.; Kinkela, T.L.; Blumberg, S.; Thomassen, H.A.; Pike, B.L.; Fair, J.N.; et al. Major Increase in Human Monkeypox Incidence 30 Years after Smallpox Vaccination Campaigns Cease in the Democratic Republic of Congo. Proc. Natl. Acad. Sci. U. S. A. 2010, 107, 16262–16267, doi:10.1073/PNAS.1005769107/SUPPL_FILE/PNAS.201005769SI.PDF.

7. Willemse, A.; Egberink, H.F. TRANSMISSION OF COWPOX VIRUS INFECTION FROM DOMESTIC CAT TO MAN. Lancet 1985, 325, 1515, doi:10.1016/S0140-6736(85)92299-8.

8. Nolen, L.D.; Osadebe, L.; Katomba, J.; Likofata, J.; Mukadi, D.; Monroe, B.; Doty, J.; Hughes, C.M.; Kabamba, J.; Malekani, J.; et al. Extended Human-to-Human Transmission during a Monkeypox Outbreak in the Democratic Republic of the Congo. Emerg. Infect. Dis. 2016, 22, 1014–1021, doi:10.3201/eid2206.150579.

9. Kantele, A.; Chickering, K.; Vapalahti, O.; Rimoin, A.W. Emerging Diseases—the Monkeypox Epidemic in the Democratic Republic of the Congo. Clin. Microbiol. Infect. 2016, 22, 658–659, doi:10.1016/J.CMI.2016.07.004.

10. Durski, K.N.; McCollum, A.M.; Nakazawa, Y.; Petersen, B.W.; Reynolds, M.G.; Briand, S.; Djingarey, M.H.; Olson, V.; Damon, I.K.; Khalakdina, A. Emergence of Monkeypox — West and Central Africa, 1970–2017. MMWR. Morb. Mortal. Wkly. Rep. 2018, 67, 306–310, doi:10.15585/mmwr.mm6710a5.

11. Hoff, N.A.; Doshi, R.H.; Colwell, B.; Kebela-Illunga, B.; Mukadi, P.; Mossoko, M.; Spencer, D.; Muyembe-Tamfum, J.-J.; Okitolonda-Wemakoy, E.; Lloyd-Smith, J.; et al. Evolution of a Disease Surveillance System: An Increase in Reporting of Human Monkeypox Disease in the Democratic Republic of the Congo, 2001–2013. Int. J. Trop. Dis. Heal. 2017, 25, 1–10, doi:10.9734/IJTDH/2017/35885.

12. Golden, J.W.; Hooper, J.W. The Strategic Use of Novel Smallpox Vaccines in the Post-Eradication World. Expert Rev. Vaccines 2011, 10, 1021–1035, doi:10.1586/erv.11.46.

13. Lane, J.M.; Goldstein, J. Adverse Events Occurring after Smallpox Vaccination. Semin. Pediatr. Infect. Dis. 2003, 14, 189–195, doi:10.1016/S1045-1870(03)00032-3.

14. Nalca, A.; Zumbrun, E.E. ACAM2000&™: The New Smallpox Vaccine for United States Strategic National Stockpile. Drug Des. Devel. Ther. 2010, 4, 71–79, doi:10.2147/DDDT.S3687.

15. Casey, C.G. Adverse Events Associated With Smallpox Vaccination in the United States, January-October 2003. JAMA 2005, 294, 2734, doi:10.1001/jama.294.21.2734.

16. Copeman, P.W.M.; Wallace, H.J. Eczema Vaccinatum. BMJ 1964, 2, 906–908, doi:10.1136/bmj.2.5414.906.

17. Kenner, J.; Cameron, F.; Empig, C.; Jobes, D. V.; Gurwith, M. LC16m8: An Attenuated Smallpox Vaccine. Vaccine 2006, 24, 7009–7022, doi:10.1016/J.VACCINE.2006.03.087.

18. Garza, N.L.; Hatkin, J.M.; Livingston, V.; Nichols, D.K.; Chaplin, P.J.; Volkmann, A.; Fisher, D.; Nalca, A. Evaluation of the Efficacy of Modified Vaccinia Ankara (MVA)/IMVAMUNE® against Aerosolized Rabbitpox Virus in a Rabbit Model. Vaccine 2009, 27, 5496, doi:10.1016/J.VACCINE.2009.06.105.

19. Hatch, G.J.; Graham, V.A.; Bewley, K.R.; Tree, J.A.; Dennis, M.; Taylor, I.; Funnell, S.G.P.; Bate, S.R.; Steeds, K.; Tipton, T.; et al. Assessment of the Protective Effect of Imvamune and Acam2000 Vaccines against Aerosolized Monkeypox Virus in Cynomolgus Macaques. J. Virol. 2013, 87, 7805–7815, doi:10.1128/JVI.03481-12.

20. Monkeypox and Smallpox Vaccine Guidance Available online: https://www.cdc.gov/poxvirus/monkeypox/clinicians/smallpox-vaccine.html.

21. Antoine, G.; Scheiflinger, F.; Dorner, F.; Falkner, F.G. The Complete Genomic Sequence of the Modified Vaccinia Ankara Strain: Comparison with Other Orthopoxviruses. Virology 1998, 244, 365–396, doi:10.1006/VIRO.1998.9123.

22. Reed, K.D.; Melski, J.W.; Graham, M.B.; Regnery, R.L.; Sotir, M.J.; Wegner, M. V; Kazmierczak, J.J.; Stratman, E.J.; Li, Y.; Fairley, J.A.; et al. The Detection of Monkeypox in Humans in the Western Hemisphere. N. Engl. J. Med. 2004, 350, 342–350, doi:10.1056/NEJMoa032299.

23. Bunge, E.M.; Hoet, B.; Chen, L.; Lienert, F.; Weidenthaler, H.; Baer, L.R.; Steffen, R. The Changing Epidemiology of Human Monkeypox-A Potential Threat? A Systematic Review. PLoS Negl. Trop. Dis. 2022, 16, e0010141, doi:10.1371/journal.pntd.0010141.

24. Multi-Country Monkeypox Outbreak: Situation Update Available online: https://www.who.int/emergencies/disease-outbreak-news/item/2022-DON396#:∼:text=Since 1 January and as,new countries have reported cases. (accessed on 15 July 2022).

25. WHO Director-General Declares the Ongoing Monkeypox Outbreak a Public Health Emergency of International Concern Available online: https://www.who.int/europe/news/item/23-07-2022-who-director-general-declares-the-ongoing-monkeypox-outbreak-a-public-health-event-of-international-concern (accessed on 24 July 2022).

26. Multi-Country Monkeypox Outbreak in Non-Endemic Countries Available online: https://www.who.int/emergencies/disease-outbreak-news/item/2022-DON385.

27. Arita, I.; Jezek, Z.; Khodakevich, L.; Ruti, K. Human Monkeypox: A Newly Emerged Orthopoxvirus Zoonosis in the Tropical Rain Forests of Africa. Am. J. Trop. Med. Hyg. 1985, 34, 781–789, doi:10.4269/ajtmh.1985.34.781.

28. Jezek, Z.; Marennikova, S.S.; Mutumbo, M.; Nakano, J.H.; Paluku, K.M.; Szczeniowski, M. Human Monkeypox: A Study of 2,510 Contacts of 214 Patients. J. Infect. Dis. 1986, 154, 551–555, doi:10.1093/infdis/154.4.551.

29. Jezek, Z.; Grab, B.; Dixon, H. Stochastic Model for Interhuman Spread of Monkeypox. Am. J. Epidemiol. 1987, 126, 1082–1092, doi:10.1093/oxfordjournals.aje.a114747.

30. Fine, P.E.; Jezek, Z.; Grab, B.; Dixon, H. The Transmission Potential of Monkeypox Virus in Human Populations. Int. J. Epidemiol. 1988, 17, 643–650, doi:10.1093/ije/17.3.643.

31. Jamieson, D.J.; Cono, J.; Richards, C.L.; Treadwell, T.A. The Role of the Obstetrician-Gynecologist in Emerging Infectious Diseases: Monkeypox and Pregnancy. Obstet. Gynecol. 2004, 103, 754–756, doi:10.1097/01.AOG.0000114987.76424.6d.

32. Huhn, G.D.; Bauer, A.M.; Yorita, K.; Graham, M.B.; Sejvar, J.; Likos, A.; Damon, I.K.; Reynolds, M.G.; Kuehnert, M.J. Clinical Characteristics of Human Monkeypox, and Risk Factors for Severe Disease. Clin. Infect. Dis. an Off. Publ. Infect. Dis. Soc. Am. 2005, 41, 1742–1751, doi:10.1086/498115.

33. Hammarlund, E.; Lewis, M.W.; Carter, S. V; Amanna, I.; Hansen, S.G.; Strelow, L.I.; Wong, S.W.; Yoshihara, P.; Hanifin, J.M.; Slifka, M.K. Multiple Diagnostic Techniques Identify Previously Vaccinated Individuals with Protective Immunity against Monkeypox. Nat. Med. 2005, 11, 1005–1011, doi:10.1038/nm1273.

34. Karem, K.L.; Reynolds, M.; Hughes, C.; Braden, Z.; Nigam, P.; Crotty, S.; Glidewell, J.; Ahmed, R.; Amara, R.; Damon, I.K. Monkeypox-Induced Immunity and Failure of Childhood Smallpox Vaccination to Provide Complete Protection. Clin. Vaccine Immunol. 2007, 14, 1318–1327, doi:10.1128/CVI.00148-07.

35. Fleischauer, A.T.; Kile, J.C.; Davidson, M.; Fischer, M.; Karem, K.L.; Teclaw, R.; Messersmith, H.; Pontones, P.; Beard, B.A.; Braden, Z.H.; et al. Evaluation of Human-to-Human Transmission of Monkeypox from Infected Patients to Health Care Workers. Clin. Infect. Dis. an Off. Publ. Infect. Dis. Soc. Am. 2005, 40, 689–694, doi:10.1086/427805.

36. Cono, J.; Cragan, J.D.; Jamieson, D.J.; Rasmussen, S.A. Prophylaxis and Treatment of Pregnant Women for Emerging Infections and Bioterrorism Emergencies. Emerg. Infect. Dis. 2006, 12, 1631–1637, doi:10.3201/eid1211.060618.

37. Kennedy, J.S.; Gurwith, M.; Dekker, C.L.; Frey, S.E.; Edwards, K.M.; Kenner, J.; Lock, M.; Empig, C.; Morikawa, S.; Saijo, M.; et al. Safety and Immunogenicity of LC16m8, an Attenuated Smallpox Vaccine in Vaccinia-Naive Adults. J. Infect. Dis. 2011, 204, 1395–1402, doi:10.1093/infdis/jir527.

38. IMVAMUNE® Smallpox Vaccine in Adult Healthcare Personnel at Risk for Monkeypox in the Democratic Republic of the Congo Available online: https://clinicaltrials.gov/ct2/show/NCT02977715?term=NCT02977715&draw=2&rank=1.

39. Centers for Disease Control and Prevention (CDC) Vaccines Available online: https://www.cdc.gov/smallpox/clinicians/vaccines.html#:∼:text=The Strategic National Stockpile (SNS,vaccines in the United States.

40. Rimoin, A.W.; Graham, B.S. Whither Monkeypox Vaccination. Vaccine 2011, 29 Suppl 4, D60–4, doi:10.1016/j.vaccine.2011.09.004.

41. Petersen, B.W.; Kabamba, J.; McCollum, A.M.; Lushima, R.S.; Wemakoy, E.O.; Muyembe Tamfum, J.-J.; Nguete, B.; Hughes, C.M.; Monroe, B.P.; Reynolds, M.G. Vaccinating against Monkeypox in the Democratic Republic of the Congo. Antiviral Res. 2019, 162, 171–177, doi:10.1016/j.antiviral.2018.11.004.

42. Harapan, H.; Wagner, A.L.; Yufika, A.; Setiawan, A.M.; Anwar, S.; Wahyuni, S.; Asrizal, F.W.; Sufri, M.R.; Putra, R.P.; Wijayanti, N.P.; et al. Acceptance and Willingness to Pay for a Hypothetical Vaccine against Monkeypox Viral Infection among Frontline Physicians: A Cross-Sectional Study in Indonesia. Vaccine 2020, 38, 6800–6806, doi:10.1016/j.vaccine.2020.08.034.

43. Yong, S.E.F.; Ng, O.T.; Ho, Z.J.M.; Mak, T.M.; Marimuthu, K.; Vasoo, S.; Yeo, T.W.; Ng, Y.K.; Cui, L.; Ferdous, Z.; et al. Imported Monkeypox, Singapore. Emerg. Infect. Dis. 2020, 26, 1826–1830, doi:10.3201/eid2608.191387.

44. The World Health Organization Operational Framework for Deployment of the World Health Organization Smallpox Vaccine Emergency Stockpile in Response to a Smallpox Event Available online: https://apps.who.int/iris/handle/10665/259574.

45. Emergent BioSolutions Awarded 10-Year HHS Contract to Deliver ACAM2000®, (Smallpox (Vaccinia) Vaccine, Live) Into the Strategic Available online: https://www.bloomberg.com/press-releases/2019-09-03/emergent-biosolutions-awarded-10-year-hhs-contract-to-deliver-acam2000-smallpox-vaccinia-vaccine-live-into-the-strategic.

46. BAVARIAN NORDIC ANNOUNCES U.S. FDA APPROVAL OF JYNNEOSTM (SMALLPOX AND MONKEYPOX VACCINE, LIVE, NON-REPLICATING) FOR PREVENTION OF SMALLPOX AND MONKEYPOX DISEASE IN ADULTS Available online: https://web.archive.org/web/20220607231106/https://www.bavarian-nordic.com/investor/news/news.aspx?news=5758.

47. HHS Orders 2.5 Million More Doses of JYNNEOS Vaccine For Monkeypox Preparedness Available online: https://web.archive.org/web/20220704005859/https://www.hhs.gov/about/news/2022/07/01/hhs-orders-2-point-5-million-more-doses-jynneos-vaccine-for-monkeypox-preparedness.html.

48. Nguyen, P.-Y.; Ajisegiri, W.S.; Costantino, V.; Chughtai, A.A.; MacIntyre, C.R. Reemergence of Human Monkeypox and Declining Population Immunity in the Context of Urbanization, Nigeria, 2017-2020. Emerg. Infect. Dis. 2021, 27, 1007–1014, doi:10.3201/eid2704.203569.

49. Bankuru, S.V.; Kossol, S.; Hou, W.; Mahmoudi, P.; Rychtář, J.; Taylor, D. A Game-Theoretic Model of Monkeypox to Assess Vaccination Strategies. PeerJ 2020, 8, e9272, doi:10.7717/peerj.9272.

50. The World Health Organization Vaccines and Immunization for Monkeypox: Interim Guidance, 14 June 2022 Available online: https://www.who.int/publications/i/item/who-mpx-immunization-2022.1.

51. Centers for Disease Control and Prevention (CDC) Smallpox/Monkeypox VIS Available online: https://www.cdc.gov/vaccines/hcp/vis/vis-statements/smallpox-monkeypox.html.

52. Centers for Disease Control and Prevention (CDC) Monkeypox and Smallpox Vaccine Guidance Available online: https://www.cdc.gov/poxvirus/monkeypox/clinicians/smallpox-vaccine.html.

53. Centers for Disease Control and Prevention (CDC) Considerations for Monkeypox Vaccination Available online: https://www.cdc.gov/poxvirus/monkeypox/considerations-for-monkeypox-vaccination.html.

54. GAVI Five Things You Need to Know about Monkeypox Available online: https://www.gavi.org/vaccineswork/five-things-you-need-know-about-monkeypox?gclid=Cj0KCQjwidSWBhDdARIsAIoTVb3syvJpkWyU3kp69Cs2ufSD0MS Rrfu5dApeh2-MkBK_hGZFrYwIqTwaAhfDEALw_wcB.

55. Teresa Wright Monkeypox: Vaccine Recommended for Canadians at High Risk of Exposure 2022.

56. The U.S. Department of Health and Human Services (HHS) HHS Announces Enhanced Strategy to Vaccinate and Protect At-Risk Individuals from the Current Monkeypox Outbreak Available online: https://www.hhs.gov/about/news/2022/06/28/hhs-announces-enhanced-strategy-vaccinate-protect-at-risk-individuals-from-current-monkeypox-outbreak.html.

57. DW Monkeypox: German Panel Recommends Vaccine for Risk Groups 2022.

58. Reuters WHO Declares Global Health Emergency over Monkeypox Outbreak 2022.

