## Supplemental Table 1 for "Vaccine Approach for Human Monkeypox over the Years and Current Recommendations to Prevent the Outbreak: A Rapid Review"

**Table 1: Summary of Included Primary Articles**

| **Author** | **Publication year** | **Article type/ Study design** | **Region/ Country** | **Vaccine Name** | **Clinical outcome of Monkeypox vaccine** |
| --- | --- | --- | --- | --- | --- |
| Arita I. et al.^(1)^ | 1985 | Assessment of special surveillance and research | Tropical rain forests of West and  Central Africa | Smallpox vaccine | A 13% case-fatality rate was found where 17 out of 131 individuals died within 3 weeks of the exposure. All of them were unvaccinated youngsters aging under seven years. The secondary attack rate among unvaccinated contacts was around 15% and was the same between 1982 and 1983 and 1970 to 1981. |
| Jezek, Z. et al.^(2)^ | 1986 | Cohort study | Zaire in the Democratic Republic of the Congo | Smallpox vaccine | The standard smallpox vaccine was 85% effective in preventing monkeypox disease. The attack rate for monkeypox among the 12,070 Unvaccinated household contacts along with smallpox infection rates was ranging from 37% to 88%. Statistical analysis of vaccination history and closeness of contact revealed that new cases of monkeypox were much more likely to occur in unvaccinated than in vaccinated contacts and in households rather than in more-casual contacts. |
| Jezek Z. et al.^(3)^ | 1987 | A computerized Monte Carlo model | Zaire, Africa | Smallpox vaccine | The secondary attack rate among the close contacts was 0.030. The attack rate was strongly related to the residence and the vaccination status of the close contacts. The secondary attack rate (for the first generation) among unvaccinated contacts who lived in the same residence as an index case was as high as 0.110. In contrast, the corresponding rate among vaccinated contacts who lived outside the affected household was almost 30 times lower (0.004). |
| Fine P.E. et al.^(4)^ | 1988 | Analysis of data on monkeypox in Zaire over the five years 1980-1984 | Zaire, Africa | Smallpox vaccine (Vaccinia) | The secondary attack rate was 0.110 for the unvaccinated contacts in the same household and 0.038 for the contacts outside the same family. The equivalent rates for contacts who had vaccinations were 0.017 and 0.004, respectively. Therefore, among those contacts with a history of vaccination—70% (1099 out of 1573) received a high level of protection from the vaccine. |
| Jezek Z. et al.^(5)^ | 1988 | Active surveillance investigation | Zaire, Africa | Smallpox vaccine | There was a significant correlation between the secondary attack rates of monkeypox and two factors: the exposed person's residence and vaccination history. It was discovered that the attack rate among individuals who had never had a vaccination (7.47%) was substantially higher than that among those who had previously received a vaccination (0.96%). Unvaccinated contacts who shared a home with a monkeypox patient had the highest attack rate (9.3%), which was seven times higher than the rate for similarly situated vaccinated household members (1.3%). |
| Jamieson D.J. et al.^(6)^ | 2004 | Commentary | USA | Smallpox vaccine (Vaccinia immune globulin) | The first evidence of community-acquired monkeypox in the United States was reported by the CDC at the beginning of June 2003. The CDC advised smallpox (vaccinia) vaccine (85% effective against monkeypox) due to the high death rate linked with monkeypox on the African continent and the lack of experience with monkeypox in the United States. Minor risk of fetal from smallpox immunization during pregnancy exists, resulting in premature birth and fetal and neonatal death. Nevertheless, given the potentially fatal risk of monkeypox infection, exposed women were urged to get the smallpox vaccine regardless of whether they were pregnant. |
| Hammarlund E. et al.^(7)^ | 2005 | Observational prospective study | USA | Smallpox vaccine (Live virus vaccine) | This study identified five vaccinated subjects who came in contact with monkeypox and three vaccinated subjects who showed complete protection against the onset of Monkeypox-induced symptoms. Although the sample size was too small to provide specific statistical estimates, the general conclusion is that approximately half of those who received the vaccine (3 of 8) continue to have long-lasting protective immunity against monkeypox. |
| Fleischauer A.T. et al.^(8)^ | 2005 | Public health investigation | USA | Smallpox vaccine | 94% of previously vaccinated, exposed HCWs tested positive for anti-orthopoxvirus IgG antibodies. No individuals appear to have had a significant change in anti-orthopoxvirus IgG levels suggestive of effects due to recent booster exposure. So, smallpox vaccination can provide protection for a long period of time. It is also unclear whether recent vaccination or infection produced a single positive IgM result; the average duration of IgM persistence after smallpox vaccination is unknown. According to anecdotal evidence with CDC vaccines, some primary vaccinees may exhibit an IgM response for up to 6 months (unpublished data). |
| Nalca A. et al.^(9)^ | 2005 | Review article | USA | Routine smallpox vaccine | Clinical sign symptoms of Monkeypox were found to be more pronounced in unvaccinated patients. Chills and/or sweats, headache, backache, sore throat, cough, shortness of breath, and lymphadenopathy have been observed in 90% of unvaccinated patients. |
| Huhn G.D. et al.^(10)^ | 2005 | Retrospective analysis of clinical reports and active and passive surveillance of suspected Monkeypox cases | USA | Smallpox vaccine | The significant finding of this study was that previous smallpox vaccination was not associated with disease severity or hospitalization. Seven patients (21%; median age: 39 years, range: 33–47 years) reported previous smallpox vaccination or had recognized smallpox scars. Nevertheless, bivariate and multivariate analyses found no difference in illness severity or inpatient hospitalization in patients with a reported history of smallpox vaccination. |
| Cono J. et al.^(11)^ | 2006 | Perspective review | USA | Smallpox vaccine (Vaccinia) | According to the US FDA's pregnancy classifications of bioterrorism medical countermeasures, the smallpox vaccine is categorized as unlicensed or category C and indicated for potential use during monkeypox infection. The use of smallpox vaccination, where they are available, for pregnant women in an emergency situation when there is a high chance of life-threatening exposure to an infectious disease will likely be advised, despite unknown dangers to the fetus. |
| Karem K.L. et al.^(12)^ | 2007 | A follow-up, household-based, case control study | USA | Smallpox vaccine | In this study, 24% of the participants had previous smallpox immunization history during childhood. The results of this study indicate that remote vaccination against smallpox (30 years earlier) does not completely protect against systemic orthopoxvirus infection; in some cases, it may prevent systemic disease, but the relative contributions of infectious inoculum and route of exposure, in addition to remote vaccination, may significantly affect whether systemic illness, asymptomatic infection or atypical illness manifest. |
| Rimoin A.W. et al.^(13)^ | 2010 | An active population-based surveillance | Democratic Republic of Congo | Smallpox vaccine | The frequency of human monkeypox has substantially increased by 20-folds in rural DRC 30 years after widespread smallpox immunization campaigns stopped. They found that the risk of human monkeypox is inversely correlated with smallpox immunization. The risk of monkeypox was 5.2 times lower in those who received vaccinations than in those who did not (0.78 vs. 4.05 per 10,000). |
| Kennedy J.S. et al.^(14)^ | 2011 | Phase I/II randomized, double-blind, comparative clinical trial | USA | LC16m8 (an attenuated cell culture–adapted Lister vaccinia smallpox vaccine) | The main objective of this trial was to compare the safety and immunogenicity of LC16m8 with Dryvax in vaccinia-naive participants. It has been found that, in order to prevent human monkeypox, LC16m8 is a feasible next-generation vaccination alternative to first-generation smallpox vaccines, at least in high-risk groups. Its clinical efficacy against human monkeypox has not yet been determined. |
| Rimoin, A.W. et al.^(15)^ | 2011 | Short commentary | Democratic Republic  of Congo | Dryvax, ACAM2000 (Live Vaccinia Vaccine) | Monkeypox risk was 5.21 times lower in vaccinated individuals compared to unvaccinated individuals, showing that >80% protection was maintained for >30 years. With an efficacy of 85% at the present incidence rate, one Monkeypox infection may be avoided for every 600 people who received the vaccine in monkeypox-endemic areas. The only smallpox vaccination that has been shown to be effective in people is live vaccine inoculation. |
| Kalthan E. et al.^(16)^ | 2018 | Monkeypox outbreak investigation Study | Alindao-Mingala  Health District of Central African Republic | Smallpox vaccine | In 87.5% of cases, the absence of smallpox vaccination was linked to severe presentations of Monkeypox. In this study, 19.2% of the participants had a smallpox vaccination scar, and the overall attack rate was lower in those who had received the vaccine (0.95/1000) compared to those who had not (3.6/1000). |
| Petersen, B.W. et al.^(17)^ | 2019 | Prospective cohort study | Democratic Republic of Congo | IMVAMUNE (Third generation smallpox vaccine) | Due to reporting so many adverse events of first- and second-generation smallpox vaccine, this study aims to follow up the cohort of health workers who have received two doses of the third-generation smallpox vaccine. Based on approaches, it was decided to study the ability of vaccination with IMVAMUNE to prevent monkeypox in DRC HCWs. The study commenced in February 2017 and is currently ongoing while study participants undergo immunologic monitoring and follow-up for exposure to monkeypox virus. |
| Harapan H. et al.^(18)^ | 2020 | Online based cross- sectional study | Indonesia | IMVAMUNE® Smallpox Vaccine | A clinical trial is ongoing to evaluate the safety and efficacy of a monkeypox vaccine among HCWs. That is why the objective of this study was to evaluate the acceptance and willingness to pay (WTP) for the vaccine among HCWs in Indonesia, where 96.0% of the participants expressed acceptance of free vaccination. The new generation of the vaccine, IMVAMUNE, has been developed with improved safety profiles, and a clinical trial is ongoing to evaluate its safety and efficacy in preventing monkeypox among HCWs in the Democratic Republic of the Congo (registered in ClinicalTrials.gov under identifier NCT02977715). |
| Yong S.E.F. et al.^(19)^ | 2020 | Case study | Singapore | Smallpox vaccination (ACAM2000; Sanofi Pasteur Biologics Co) | In May 2019, a traveler from Nigeria to Singapore was investigated as a confirmed monkeypox case. 8 lower risk contacts and 23 close contacts were found. Smallpox vaccination was made available to close contacts as postexposure prophylaxis. Of the 22 close contacts, 14 chose to receive the immunization, 2 had contraindications, and 6 contacts refused to get vaccinated. On days 6–8 of the review, a scab or ulcer was present in every vaccine recipient. Slight fever and minor swelling at the injection site were side effects, but no serious adverse events were reported. |
| Bankuru S.V. et al.^(20)^ | 2020 | Compartmental epidemiological model, game theory approach | Worldwide | Smallpox vaccine | To evaluate vaccination decision-making, a game-theoretical approach was used. The model quantifies the smallpox vaccine's costs and advantages. This study determined that the ideal vaccination rate is approximately 0.04, meaning people should get vaccinated once every 25 years. Additionally, they discovered that monkeypox disease is preventable and can be eliminated through vaccination in a semi-endemic equilibrium. However, vaccination alone cannot wholly eradicate monkeypox in an equilibrium where it is entirely endemic. |
| Whitehouse E. R. et al.^(21)^ | 2021 | Surveillance | Democratic Republic of the Congo | Smallpox vaccine | The incidence among confirmed case patients was nearly three times greater (16.4 per 100 000) among those assumed to be unvaccinated than those assumed to be vaccinated (6.0 per 100 000). The incidence among those who were assumed to have had vaccinations was similar to that in the Bumba zone between 1981 and 1985 (6.3 per 100 000). These results support earlier studies that suggested a degree of cross-protection against monkeypox might be conferred by historical vaccination against smallpox. |
| Nguyen, P.Y. et al.^(22)^ | 2021 | Review of retrieved epidemiologic and demographic data from monthly and weekly epidemiologic reports | Nigeria | Smallpox vaccine | Only 10.1% of Nigeria's population had received the smallpox vaccine as of 2016, and the serologic immunity level was 25.7% among those who had received the vaccination compared to 2.6% in the general population. Using the expected linear rate of decline over time from vaccination, the cross-immunity protection for monkeypox of 85% of smallpox vaccination decreased to just 23.1% among those who received it. Epidemiological data suggest that having received a smallpox vaccination in the past offers at least some protection against serious monkeypox infections. The total population immunity level has significantly decreased during the previous 45 years due to population expansion in the postvaccination era. So, the role of vaccination in preventing monkeypox is being considered, and clinical trials for healthcare workers have been suggested here. |

References:

1.	Arita I, Jezek Z, Khodakevich L, Ruti K. Human monkeypox: a newly emerged orthopoxvirus zoonosis in the tropical rain forests of Africa. Am J Trop Med Hyg. 1985 Jul;34(4):781–9.

2.	Jezek Z, Marennikova SS, Mutumbo M, Nakano JH, Paluku KM, Szczeniowski M. Human monkeypox: a study of 2,510 contacts of 214 patients. J Infect Dis. 1986 Oct;154(4):551–5.

3.	Jezek Z, Grab B, Dixon H. Stochastic model for interhuman spread of monkeypox. Am J Epidemiol. 1987 Dec;126(6):1082–92.

4.	Fine PE, Jezek Z, Grab B, Dixon H. The transmission potential of monkeypox virus in human populations. Int J Epidemiol. 1988 Sep;17(3):643–50.

5.	Jezek Z, Grab B, Szczeniowski M V, Paluku KM, Mutombo M. Human monkeypox: secondary attack rates. Bull World Health Organ. 1988;66(4):465–70.

6.	Jamieson DJ, Cono J, Richards CL, Treadwell TA. The role of the obstetrician-gynecologist in emerging infectious diseases: monkeypox and pregnancy. Obstet Gynecol. 2004 Apr;103(4):754–6.

7.	Hammarlund E, Lewis MW, Carter S V, Amanna I, Hansen SG, Strelow LI, et al. Multiple diagnostic techniques identify previously vaccinated individuals with protective immunity against monkeypox. Nat Med. 2005 Sep;11(9):1005–11.

8.	Fleischauer AT, Kile JC, Davidson M, Fischer M, Karem KL, Teclaw R, et al. Evaluation of human-to-human transmission of monkeypox from infected patients to health care workers. Clin Infect Dis an Off Publ Infect Dis Soc Am. 2005 Mar;40(5):689–94.

9.	Nalca A, Rimoin AW, Bavari S, Whitehouse CA. Reemergence of monkeypox: prevalence, diagnostics, and countermeasures. Clin Infect Dis an Off Publ Infect Dis Soc Am. 2005 Dec;41(12):1765–71.

10.	Huhn GD, Bauer AM, Yorita K, Graham MB, Sejvar J, Likos A, et al. Clinical characteristics of human monkeypox, and risk factors for severe disease. Clin Infect Dis an Off Publ Infect Dis Soc Am. 2005 Dec;41(12):1742–51.

11.	Cono J, Cragan JD, Jamieson DJ, Rasmussen SA. Prophylaxis and treatment of pregnant women for emerging infections and bioterrorism emergencies. Emerg Infect Dis. 2006 Nov;12(11):1631–7.

12.	Karem KL, Reynolds M, Hughes C, Braden Z, Nigam P, Crotty S, et al. Monkeypox-induced immunity and failure of childhood smallpox vaccination to provide complete protection. Clin Vaccine Immunol. 2007 Oct;14(10):1318–27.

13.	Rimoin AW, Mulembakani PM, Johnston SC, Lloyd Smith JO, Kisalu NK, Kinkela TL, et al. Major increase in human monkeypox incidence 30 years after smallpox vaccination campaigns cease in the Democratic Republic of Congo. Proc Natl Acad Sci U S A. 2010 Sep;107(37):16262–7.

14.	Kennedy JS, Gurwith M, Dekker CL, Frey SE, Edwards KM, Kenner J, et al. Safety and immunogenicity of LC16m8, an attenuated smallpox vaccine in vaccinia-naive adults. J Infect Dis. 2011 Nov;204(9):1395–402.

15.	Rimoin AW, Graham BS. Whither monkeypox vaccination. Vaccine. 2011 Dec;29 Suppl 4(Suppl 4):D60-4.

16.	Kalthan E, Tenguere J, Ndjapou SG, Koyazengbe TA, Mbomba J, Marada RM, et al. Investigation of an outbreak of monkeypox in an area occupied by armed groups, Central African Republic. Med Mal Infect. 2018 Jun;48(4):263–8.

17.	Petersen BW, Kabamba J, McCollum AM, Lushima RS, Wemakoy EO, Muyembe Tamfum J-J, et al. Vaccinating against monkeypox in the Democratic Republic of the Congo. Antiviral Res. 2019 Feb;162:171–7.

18.	Harapan H, Wagner AL, Yufika A, Setiawan AM, Anwar S, Wahyuni S, et al. Acceptance and willingness to pay for a hypothetical vaccine against monkeypox viral infection among frontline physicians: A cross-sectional study in Indonesia. Vaccine. 2020 Oct;38(43):6800–6.

19.	Yong SEF, Ng OT, Ho ZJM, Mak TM, Marimuthu K, Vasoo S, et al. Imported Monkeypox, Singapore. Emerg Infect Dis. 2020 Aug;26(8):1826–30.

20.	Bankuru SV, Kossol S, Hou W, Mahmoudi P, Rychtář J, Taylor D. A game-theoretic model of Monkeypox to assess vaccination strategies. PeerJ. 2020;8:e9272.

21.	Whitehouse ER, Bonwitt J, Hughes CM, Lushima RS, Likafi T, Nguete B, et al. Clinical and Epidemiological Findings from Enhanced Monkeypox Surveillance in Tshuapa Province, Democratic Republic of the Congo During 2011-2015. J Infect Dis. 2021 Jun;223(11):1870–8.

22.	Nguyen P-Y, Ajisegiri WS, Costantino V, Chughtai AA, MacIntyre CR. Reemergence of Human Monkeypox and Declining Population Immunity in the Context of Urbanization, Nigeria, 2017-2020. Emerg Infect Dis. 2021 Apr;27(4):1007–14.
