## Supplemental Table 2 for "Vaccine Approach for Human Monkeypox over the Years and Current Recommendations to Prevent the Outbreak: A Rapid Review"

**Table 2: Most recent vaccine recommendations by different organizations.**

| **Article** | **Recommended by** | **Time and Date** | **Vaccine Recommendations** |
| --- | --- | --- | --- |
| [Vaccines and immunization for monkeypox: Interim guidance](https://www.who.int/publications/i/item/who-mpx-immunization-2022.1) | World Health Organization (WHO) | 14 June 2022 | Mass vaccination for monkeypox is not required. It is advised to administer a suitable second- or third-generation vaccine to contacts of patients as post-exposure prophylaxis, ideally within four days of the initial exposure. Pre-exposure prophylaxis is advised for healthcare workers at risk, lab employees who handle orthopoxviruses, and clinical laboratory staff who do monkeypox diagnostic tests. A robust information campaign, a solid pharmacovigilance program, and joint vaccine effectiveness studies with standardized methodologies and data gathering technologies are all required to support vaccination programs. Smallpox or monkeypox vaccination decisions should be based on a thorough analysis of risks and benefits on a case-by-case basis. |
| [Smallpox/Monkeypox Vaccine (JYNNEOS™): What You Need to Know](https://www.cdc.gov/vaccines/hcp/vis/vis-statements/smallpox-monkeypox.html) | Centers for Disease Control and Prevention (CDC) | 01 June 2022 | JYNNEOS™ vaccine is approved by the Food and Drug Administration (FDA) to prevent monkeypox disease in adults 18 years or older at high risk for monkeypox infection. CDC recommends JYNNEOS™ for certain laboratory workers, clinic teams who care for patients infected with orthopoxvirus, and emergency response team members who might be exposed to the viruses. JYNNEOS™ is usually 2 doses, 4 weeks apart. People who have received other types of smallpox vaccine in the past might need 1 dose only. Booster doses are recommended every 2 or 10 years if a person remains at continued risk for exposure to orthopoxviruses. It has been recommended to receive JYNNEOS™ due to exposure to the monkeypox virus regardless of concurrent illnesses, pregnancy, breastfeeding, or weakened immune system. |
| [Monkeypox and Smallpox Vaccine Guidance](https://www.cdc.gov/poxvirus/monkeypox/clinicians/smallpox-vaccine.html) | Centers for Disease Control and Prevention (CDC) | 02 June 2022 | In conjunction with the Advisory Committee on Immunization Practices (ACIP), the CDC provided recommendations on who should receive smallpox vaccination (JYNNEOS/ ACAM2000) in a non-emergency setting. At the time, ACAM2000 immunization was advised for military personnel and lab workers who handled specific orthopoxviruses. ACAM2000 vaccination has the potential for more side effects and adverse events than the newer vaccine, JYNNEOS. Thus, on November 3, 2021, ACIP recommended JYNNEOS pre-exposure prophylaxis as an alternative to ACAM2000 for some people at risk of orthopoxviruses. In order to prevent monkeypox infection, the CDC advises that the vaccination be administered within 4 days of the date of exposure. A vaccine administered between 4 and 14 days after exposure may not prevent the disease, but it may lessen the symptoms. People exposed to the monkeypox virus and who have not had the smallpox vaccine within the last three years should think about receiving it. |
| [Considerations for Monkeypox Vaccination](https://www.cdc.gov/poxvirus/monkeypox/considerations-for-monkeypox-vaccination.html) | Centers for Disease Control and Prevention (CDC) | 30 June 2022 | Currently, JYNNEOS (also known as Imvamune or Imvanex) and ACAM2000, two vaccines approved by the U.S. Food and Drug Administration (FDA), are accessible to prevent monkeypox infection. There is currently no information on these vaccinations' effectiveness in the ongoing outbreak. ACAM2000 vaccination should not be administered to those with certain medical issues, such as weakened immune system (e.g., HIV), cardiac disease, eye disease treated with topical steroids, congenital or acquired immune deficiency disorders, atopic dermatitis/eczema, infants, or pregnancy. The Advisory Committee on Immunization Practices (ACIP) decided on November 3, 2021, to suggest JYNNEOS pre-exposure prophylaxis as a substitute for ACAM2000 for some people who may be exposed to orthopoxviruses. |
| [Five things you need to know about monkeypox](https://www.gavi.org/vaccineswork/five-things-you-need-know-about-monkeypox?gclid=Cj0KCQjwidSWBhDdARIsAIoTVb3syvJpkWyU3kp69Cs2ufSD0MSRrfu5dApeh2-MkBK_hGZFrYwIqTwaAhfDEALw_wcB) | Gavi, The Vaccine Alliance (GAVI) | 17 May 2022 | The smallpox vaccine was vital to eradicating smallpox decades ago, and this vaccine can be highly effective – 85% – in preventing monkeypox. However, first-generation smallpox vaccines are no longer offered to the general population. For the protection of smallpox and monkeypox, a more recent vaccination based on vaccinia was licensed in 2019; however, it is also not yet widely accessible. |
| [Monkeypox: Vaccine recommended for Canadians at high risk of exposure](https://globalnews.ca/news/8912191/monkeypox-vaccine-recommended-canadians-exposure-phac/?fbclid=IwAR3LWRSstL-NyBG2sqLA28ychCkqI8X5ayP1Bu9e2JB6pHk1zHkTKWyMPRA) | National Advisory Committee on Immunization (NACI)-Canada | 10 June 2022 | Health Canada has authorized Imvamune; a vaccine often used to treat smallpox and monkeypox. Anyone who has come into touch with a case or has been in an environment with a high chance of exposure is given one dosage. Additionally, a second dose was advised to be given under specific conditions only. Immunocompromised individuals, pregnant or nursing women, as well as children and young people, may be administered vaccinations if their risk of exposure is higher. Given the scope of the outbreaks so far, mass vaccination campaigns against monkeypox among the populace are not currently required due to the size of the outbreaks. |
| [HHS Announces Enhanced Strategy to Vaccinate and Protect At-Risk Individuals from the Current Monkeypox Outbreak](https://www.hhs.gov/about/news/2022/06/28/hhs-announces-enhanced-strategy-vaccinate-protect-at-risk-individuals-from-current-monkeypox-outbreak.html) | The U.S. Department of Health and Human Services (HHS) | 28 June 2022 | In an effort to stop the spread of monkeypox, the U.S. Department of Health and Human Services (HHS) unveiled an improved national vaccination program. The plan will vaccinate and safeguard persons susceptible to monkeypox, prioritize vaccine distribution in areas with the greatest caseloads, and offer direction to state, territorial, tribal, and municipal health officials to help with their preparation and response operations. A four-tier distribution plan for the JYNNEOS vaccine will be used, giving priority to regions with the most significant prevalence of monkeypox cases. The number of people at risk for monkeypox who also have pre-existing illnesses, such as HIV, will determine how many doses of JYNNEOS are distributed within each tier. In order to guarantee that the communities with the greatest need have access to immunizations to prevent monkeypox, HHS will continue to develop and strengthen its vaccine supply and distribution strategy. |
| [Monkeypox: German panel recommends vaccine for risk groups](https://www.dw.com/en/monkeypox-german-panel-recommends-vaccine-for-risk-groups/a-62084728?fbclid=IwAR3D0mE6byJ768HJpTLwFxs39tM4IV4YrKw8rSQniTrfBqc2Sy_CXwEOFf8) | Germany's Standing Committee on Vaccination (STIKO) | 09 June 2022 | The vaccine advisory body advised Imvanex smallpox vaccine from Bavarian Nordic. The panel also recommended that because vaccine supplies are limited, those who have recently been exposed to the monkeypox virus should be the first to receive it. According to STIKO, the recommended vaccination course with Imvanex entails two doses given at least 28 days apart to individuals who have never received a smallpox vaccination and one dose for those who have. |
